# The Prevention and Control of Cancer by Metformin in Patients with Type 2 Diabetes: A Systematic Mapping Review

**DOI:** 10.1101/2021.06.04.21258310

**Authors:** Albania Mitchell, Michelle Price, Gabriela Cipriano

## Abstract

**Objective:** Metformin is commonly used as a first line therapy for type 2 diabetes; however, existing evidence suggests an influence in oncology. The objective of this systematic mapping review was to describe current literature regarding metformin and its role in preventing and /or controlling cancer in patients with type 2 diabetes.

**Method:** We searched PubMed, Cochrane Library, and ClinicalTrials.gov in February 2018 and April 2019 to identify research studies, systematic reviews and meta-analyses. Of the 318 citations identified, 156 publications were included in this analysis.

**Results:** The most common cancer types researched were colorectal, liver, prostate, lung and breast with the United States contributing the most to this data. Author teams averaged six members and most studies were funded. Only 68% of the articles were available open access. Ovarian and esophageal cancers were amongst the least studied, but the most costly for care.

## Introduction

Cancer is a compilation of related disease states that can exist as hematologic or solid tumors; and can range from being benign to malignant. Cancer occurs when a cell abnormally and exponentially proliferates, and/or when apoptosis is inhibited. Approximately, 34.4% of the population will have cancer at some point in their lifetime. Cancer is the second most common cause of death in the United States. Mortality is higher amongst men compared to women (196.8 per 100,000) with the highest prevalence being amongst African American men (239.9 per 100,000). In 2017, national spending for cancer care in the United States was $147.3 billion and is expected to increase over time.^1^

Diabetes is primarily characterized by symptoms of hyperglycemia. Type 2 diabetes, the most common type, occurs when the body does not properly utilize insulin to maintain normal blood glucose. Hemoglobin A1c (HgbA1c), fasting plasma glucose, and an oral glucose tolerance test are diagnostic tools used to confirm diagnosis of diabetes in the absence of hyperglycemia symptoms.^2^ According to the Centers for Disease Control and Prevention National Diabetes Statistics Report, in 2020, 34.2 million Americans have diabetes. The diagnosis of diabetes affects all facets of life. The average medical expenditure for patients with diabetes is more than double of those without diabetes.^3^

Inadequate treatment and management of this chronic disease state can lead to a plethora of life altering micro-and macro-vascular complications, such as retinopathy, nephropathy, neuropathy, and cardiovascular disease. Inadequate perfusion often contributes to poor wound healing, foot complications, and infection. Patients with diabetes are more likely to have various cardiovascular complications, such as an increased risk of high blood pressure, heart disease and stroke. Severe uncontrolled diabetes is the leading cause of adult blindness, kidney failure and non-traumatic amputations.^4^

Patients with Type 2 diabetes are at an increased risk of liver, pancreas, endometrium, colon/rectum, breast, and bladder cancer. Conversely, diabetes is also correlated with a reduced risk of prostate cancer.^5^ Both disease states, cancer and diabetes, share common risk factors like aging, obesity, diet and a sedentary lifestyle. Although this is still a developing area of research, possible ways that diabetes has an influence on cancer prevalence is hyperinsulinemia, hyperglycemia, and inflammation.^6^ A study conducted by Ranc et al. concluded that pre-existing diabetes at the time of cancer diagnosis increases mortality, compared with cancer patients without diabetes.^7^

Metformin is a biguanide antidiabetic agent currently recommended as the first-line agent to treat type 2 diabetes mellitus (T2DM) according to the American Diabetes Association.^2^ Metformin, unlike most other antidiabetic drugs, does not increase insulin secretion to decrease blood glucose levels. Through the activation of the activated protein kinase, 5’ AMP-activated protein kinase, metformin is able to decrease hepatic glucose production, decrease intestinal absorption of glucose, and improve insulin sensitivity by increasing peripheral glucose uptake and utilization. This reduces the risk of hyperinsulinemia, which is thought to increase cancer risk among diabetic patients. Metformin’s ability to lower blood glucose without causing hypoglycemia, unlike sulfonylureas, in combination with its affordable price makes it a favored choice amongst healthcare professionals and patients living with diabetes. It is fairly tolerable, with the most common side effect being dose-dependent gastrointestinal disturbances.^8^

A mapping review categorizes existing literature on a specific topic. It is used to inform policy-makers, funders and researchers on the quantity of literature existing to address a topic, as well as, illuminate variables such as population size, study design, and location. The resulting data can inform researchers of the subsets of data available for further review, including those topics with enough homogenous data for a meta-analysis. Finally, a systematic mapping review reveals popular subtopics and data gaps to allow for prioritized funding based on evidence.^9,10^

Both cancer and diabetes have been known to decrease life expectancy while increasing medical expenditures. The use of metformin in diabetes has been revolutionary in providing a medication with recognized efficacy, small side effect profile, and a minimal financial burden. Metformin and its use in cancer could improve patient outcomes in those with or at risk of both disease states.^5^ This review is intended to be used as a tool for clinical application and a stepping stone for further research. It will contribute to providers by supplying key information needed in order to make evidence-based clinical decisions in the intervention of metformin in a cancer and diabetic medical regimen. Information present in this review is also crucial in deciding where investments will be allocated in primary research in order to fill gaps in knowledge, or continue further development in areas of interest.^11^

## Materials and Methods

At the time of publication, PROSPERO, an international registry of Systematic Reviews, was not accepting submissions of Systematic Mapping Reviews. Therefore, the protocol for this review, although created according to the Prospero Guidelines^12^, was published in an institutional repository.^13^ We performed a systematic mapping review and the reporting is in accordance with the Preferred Reporting Items for Systematic Reviews and Meta-Analysis (PRISMA) guidelines.^14^

### Search Strategy

The following databases were searched for all available dates, PubMed, Cochrane Library (Cochrane Database of Systematic Reviews, Database of Abstracts of Reviews of Effects, Cochrane Central Register of Controlled Trials, Cochrane Methodology Register, Health Technology Assessment Database, NHS Economic Evaluation Database) and Clinicaltrials.gov. The initial search was performed in February 2018. Separate search strategies were created for each database based on the search terms diabetes mellitus type 2, cancer and metformin. An update on the search was conducted in October of 2019. At that time Cochrane library had updated its databases to include only the Cochrane Database of Systematic Reviews, and the Cochrane Central Register of Controlled Trials.^15^ Additionally, Cochrane Central began to include clinical trial information from The World Health Organization’s International Clinical Trials Registry Platform (ICTRP) in April of 2019.^16^ While scoping the research question, it was discovered that Chin-Hsiao Tseng from the National Taiwan University College of Medicine, has produced several published articles on the use of metformin and cancer. The full bibliography of works as listed in the ORCID profile was searched in October 2019.^17^ All PubMed searches were completed with legacy PubMed. (Appendix 1).

### Study Selection

Research studies, including systematic reviews and meta-analysis, were included. Non-systematic reviews were excluded. There were no restrictions on the date of publication. Articles published in English and Spanish were considered. Adult patients with or without cancer that had type 2 diabetes were the target population. The study had to include metformin as an intervention and address the prevention or control of cancer.

Interrater reliability was established by conducting a review of five articles using the established inclusion and exclusion criteria prior to the study screening (GC, AM). All titles and abstracts were then assessed independently to identify articles requiring full-text review (GC, AM). Any discrepancies were resolved by the third author (MP). Two reviewers assessed the full-text of the articles (GC, AM) and the third author again resolved discrepancies (MP). This same procedure was completed for the updated search.

## Results

We identified 318 citations, 263 from the first search and 55 from the updated search. Seven duplicates were removed and 311 titles and abstracts were screened. There were 209 articles excluded and 102 reviewed in full-text for eligibility. During the full-text screening, 40 articles were excluded; 24 were the wrong study type, six were not available in English or Spanish, five articles contained duplicate studies, two did not have metformin as the intervention, one was an ongoing clinical trial, one did not have cancer as an outcome, and one did not have the prevention or control of cancer. From the included references, 94 additional articles were identified; 75 from the first search and 19 from the updated search. A total of 156 articles were included in the analysis (figure 1).

**Figure 1.**
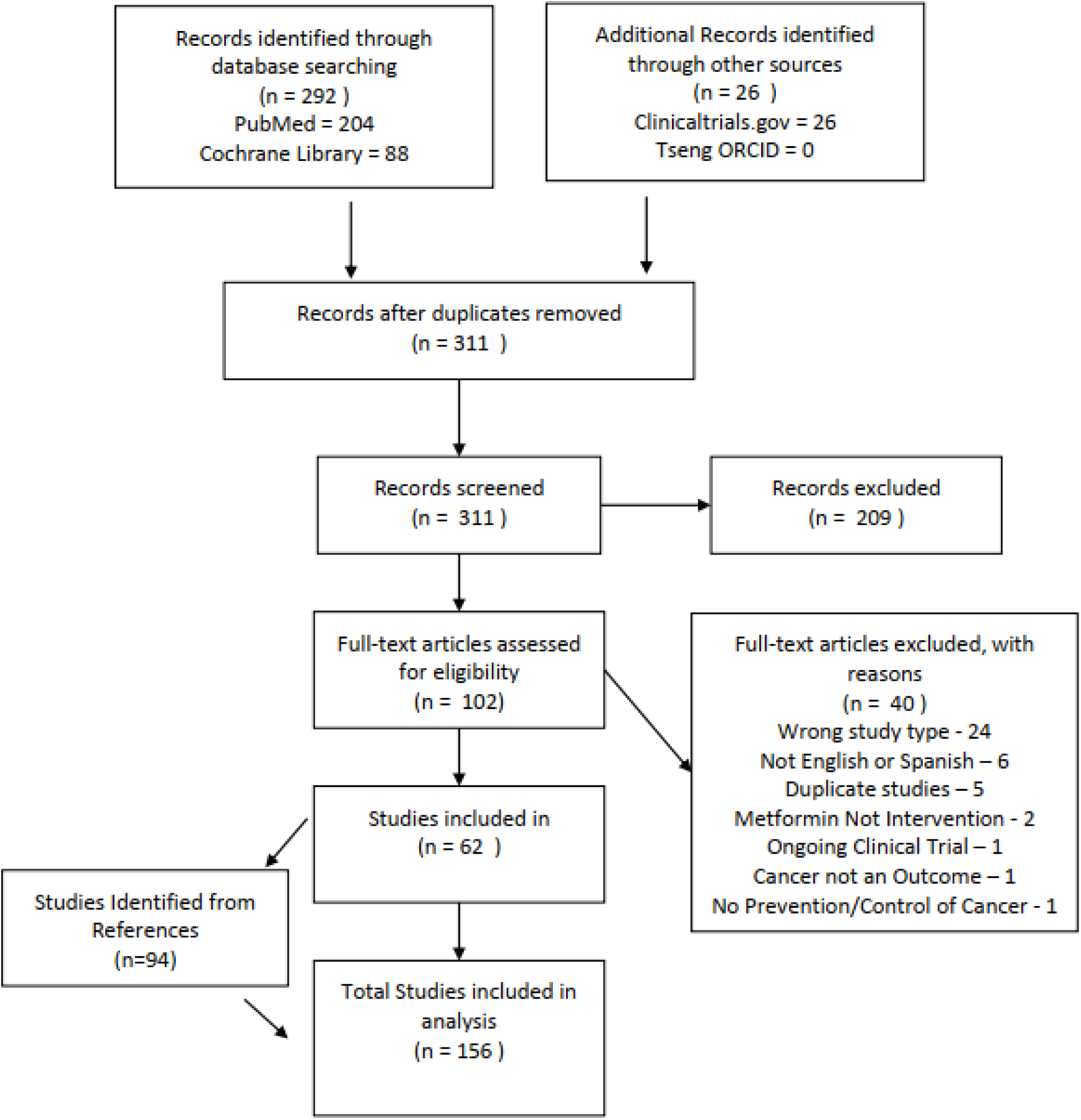
PRISMA flow chart for study selection Study Characteristics.

Of the 156 articles, there was a mixture of study types; 76 were retrospective cohorts^5,18–92^, 27 were case-control studies^93–119^, 15 were meta-analyses^120–134^, 15 were prospective cohorts^135–149^, 11 were systematic reviews with a meta-analysis^150–160^, seven were nested case-control^161–167^, two were randomized control studies^168–169^, two were systematic reviews^170–171^, and one was a cross-sectional study^172^.

These study articles were analyzed for various study characteristics, such as number of authors identified in each article, study type and number of participants (table 1), as well as the type or title of the journal the study was published in. For the 156 studies included in the data extraction and analysis, the number of authors for each of these articles ranged from 1-21. The included articles had both an average and median of six authors. The systematic reviews and meta-analysis study types, yielded the same number of authors including a mode of 5 for all study type data sets. The number of participants in each study were also included and extracted by study type. Of the 156 studies, the retrospective and prospective cohorts had the highest number of participants with an average of 127,466 and 60,061 participants, respectively. Survival analysis or time to event data had the least number of participants with an average of 1131. The average number of studies included in the systematic reviews, 32, was about doubled that of the meta-analysis, 14 (Table 1).

**Table 1:** Study Details: number of authors, number of participants, number of included studies.

| **Study Characteristic** | **Mean** | **Median** |
| --- | --- | --- |
| **Number of Authors** |  |  |
| All Articles | 6 | 6 |
| Meta Analysis, Systematic Reviews,<br>Systematic Review + Meta-Analysis | 6 | 6 |
| **Number of Participants** |  |  |
| Case control | 25,670 | 2,763 |
| Cross sectional | 14,345 | 14,345 |
| Nested Case-Control | 12,332 | 1,700 |
| Prospective | 52,301 | 2,924 |
| Retrospective | 125,792 | 27,805 |
| Time to event | 1,131 | 974 |
| **Number of Included Studies** |  |  |
| Meta-Analysis | 14 | 11 |
| Systematic Review | 32 | 32 |
| Systematic Review + Meta-Analysis | 17 | 14 |

### Cancer Type

Of the 156 articles, 115 were about a single cancer, 41 were about various cancers and seven articles did not specify in any determinable way. Of the 115 single cancer studies, there were 25 colorectal, 17 liver, 14 prostate, 14 lung, 14 breast, six pancreatic, four ovarian, three endometrial, two esophageal, two gastric, and only one each for bladder, cervical, intrahepatic cholangiocarcinoma, kidney, laryngeal, nasopharyngeal, oral, skin and thyroid (figure 2). There were 40 studies that addressed various cancer types. The following is the number of occurrences of each type in those articles; colorectal 28, breast 26, any/all cancer types 25, pancreatic 24, prostate 24, liver 20, lung 18, esophageal seven, bladder six, Ovarian six, Gastric five, Skin five, Cervical four, endometrial three, laryngeal two, oral two, Nasopharyngeal one, and thyroid had one occurrence (figure 3).

**Figure 2:**
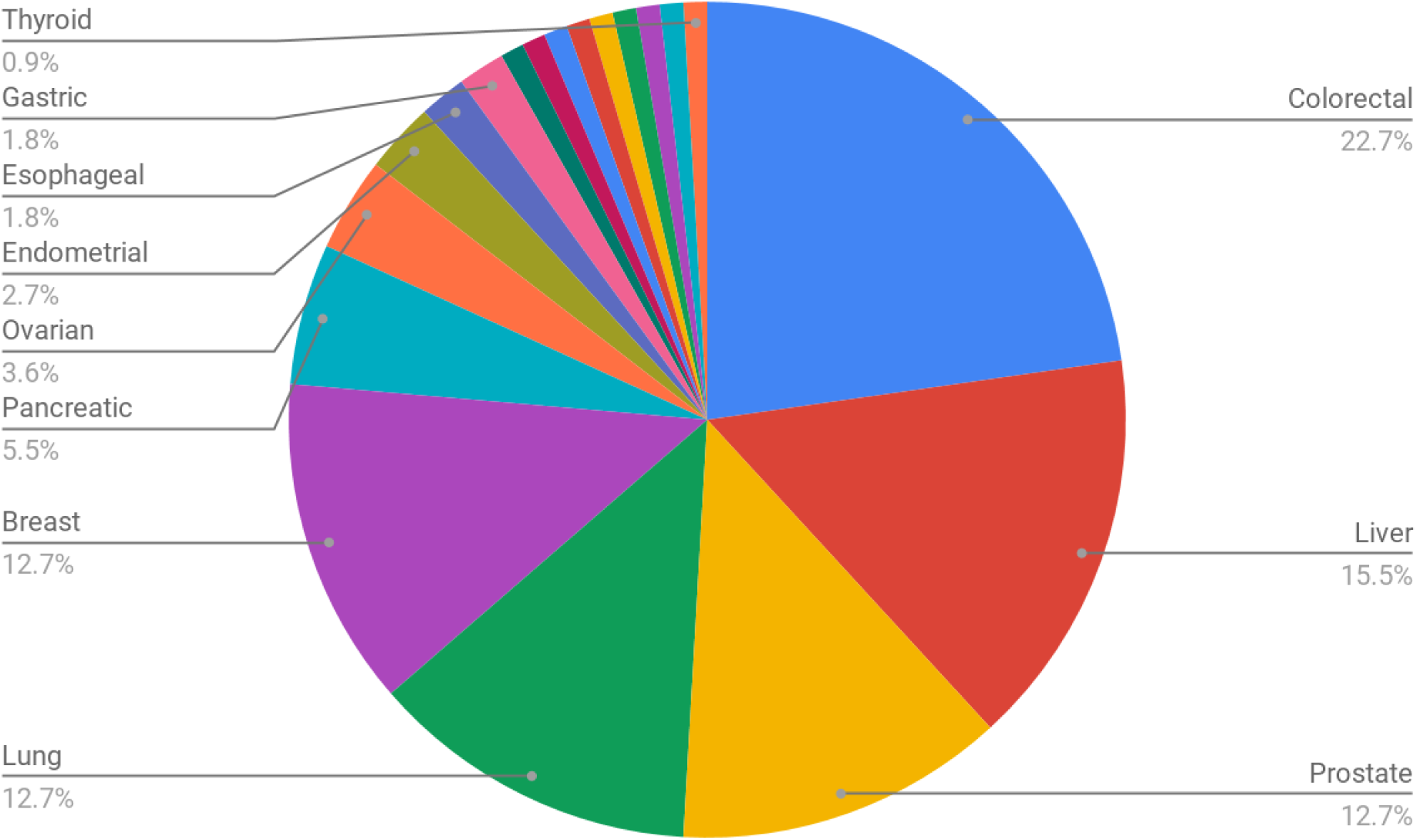
Cancer Type in studies that addressed only one type of cancer.

**Figure 3:**
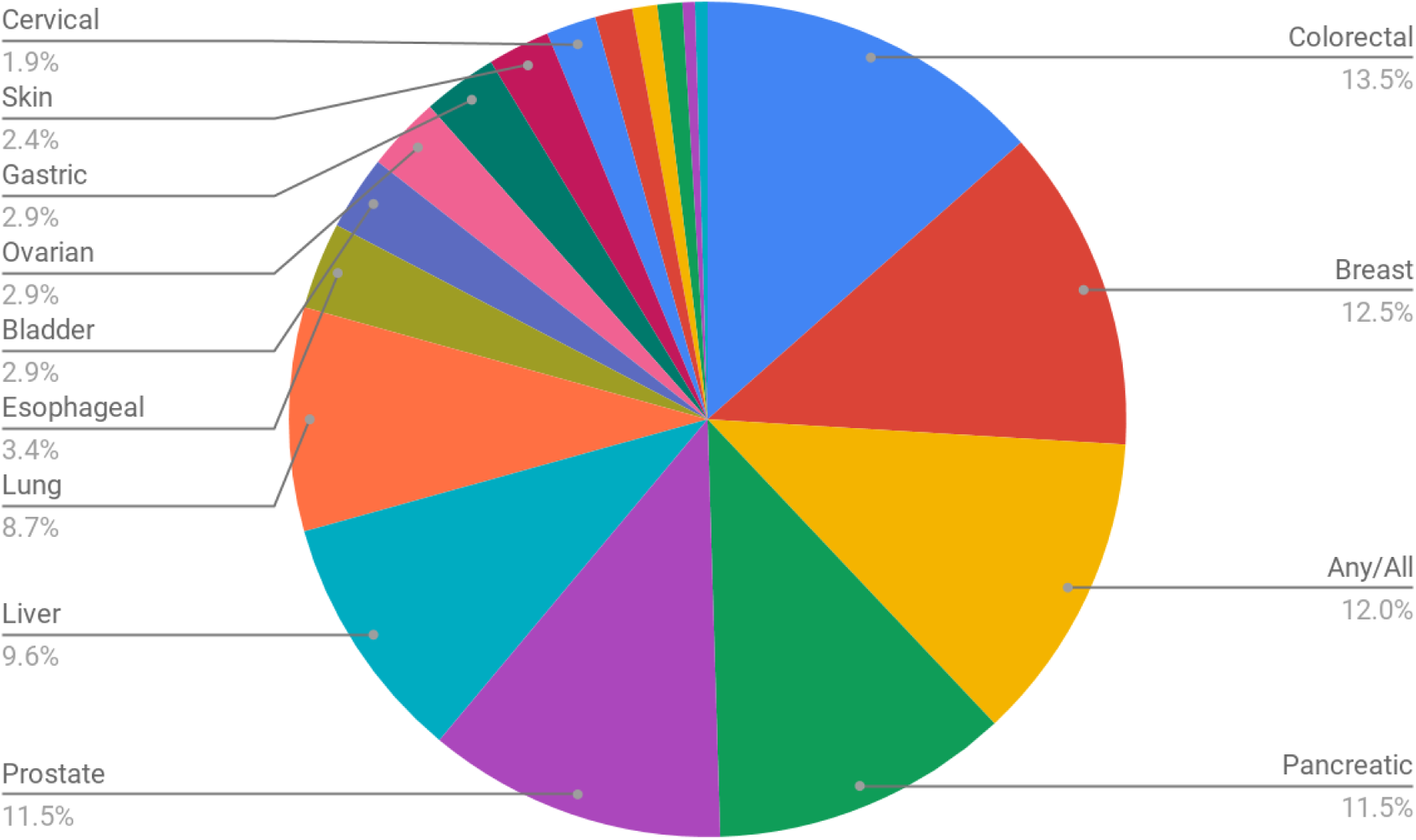
Cancer types in studies that addressed various cancers.

### Funding

For the purposes of our review, funding is defined as monetary and non-financial support of research. A majority of the studies had at least one (mode) source of funding, 67% (104 out of 156). The average number of funding sources for studies funded was 2.34. The highest amount of funding for a single study was nine sources. This was for a case-control study out of Finland about prostate cancer^115^.

Out of the 104 studies that were funded, 28 were funded by organizations based out of the United States, almost always to use data from the United States. Hagberg^163^ was the exception. This study received funding from the Intramural Research Program of the National Institutes of Health, National Cancer Institute to analyze data from the United Kingdom (UK) Clinical Practice Research Datalink (CPRD). Of the 28 studies that were funded out of the United States, 22 of them were funded by the National Institute of Health.^18,20,21,42,56,62,64,91,101,103–107,109,111,116,119,129,137,147,163^.

This systematic mapping review analyzed the common study types that were published about this topic, this is described elsewhere. The authors also took into consideration that the type of study may have impacted funding. All study types with more than one article published were included in this analysis. The highest average of funding sources belonged to nested case control (2.57 funding sources), case control (1.96 funding sources) and retrospective cohort (1.9 funding sources). The lowest average of funding sources belonged to systematic reviews and time-to-event studies. Both averaged 0.5 of funding sources.

Diabetes is known to cause an increased risk in certain cancer types, such as bladder, breast, endometrial, liver, and pancreatic cancer. Conversely, it can also cause a decreased risk of prostate cancer. Taking that into consideration it is important to analyze funding sources under the lense of cancer type.

The most common cancer types are prostate, lung, and colon cancer^173^. This is reflected in the studies that were included the most in this systematic mapping review. The studies that were published the most pertained to colorectal cancer (25), liver (17), lung (14), prostate (14) and breast cancer (13). Each of those received funding 64%, 65%, 50%, 93%, and 54% of the time respectively. Although the one study on thyroid cancer was funded, amongst cancers with more than one publication, prostate cancer was the most frequently funded. The cancer types that only contributed one study each to this systematic mapping review includes bladder, cervical, kidney, laryngeal, oropharyngeal, oral, skin and thyroid cancer. All of these studies received one source of funding except laryngeal and skin cancer.

### Country of Data Source

To summarize the origin source of the data that was analyzed during this systematic mapping review we included all literature that was not a systematic review or meta analysis. This resulted in a total of 129 articles out of 156. Overall, three countries contributed the most literature about metformin and its impact on cancer. The top three countries with the most publishings were the United States (35 or 27%), ^18–21,33,40,42,44,53,54,56,58,62,64–66,69,90,91,101,103–107,109,111,113,116–119,136,137,147^ Taiwan (30 or 23%), ^25,26,36,45,47,49,50,71–87,102,135,142,145,162,167^ and the United Kingdom (21 or 16%) ^29–32,37,48,60,88,89,92–98,112,161,163,165,166^. It is interesting to note that over 50% of the literature published that originated in Taiwan came from one single author, Tseng^17^, over the span of six years (2012 to 2018).

### Journals and Open Access

There were several journals that had publications on the topic of metformin use and its association with cancer outcomes. Of the 156 articles that data was extracted from for our analysis, *Diabetes Care* had the highest number of publications,16, regarding metformin use and cancer occurence or outcomes, followed by *Cancer Epidemiology, Biomarkers and Prevention*, which had eight. The majority of the journals had only one published article describing findings relating to metformin use and cancer (see Appendix 3).

There were 74 articles accessible via PubMed Central, which accounts for 47% of the total articles. There were 100 articles directly accessible via open access on the Publisher’s website, or 64% of the total. Combining the two methods of open access, 106 or 68% of the total articles were available open access, the rest remained behind a paywall. There were 25 instances, where articles were open access on the publisher’s website, but not available in PubMed Central. Of those 25 instances, there were four journals that were represented at least twice: American Journal of Epidemiology; Cancer, Epidemiology, Biomarkers, & Prevention; Diabetes Care; and Diabetologia.

### Publication Dates

The publication dates of the articles ranged from 2004-2018, the bulk being published between 2011 and 2014. From 2004-2009, there were only articles published using data from the United States. Continuing from 2010 to present, the United States represented at least 60% of the total publications, except for in 2010, when the percentage contributed by the United States dropped to fifty. The first systematic review or meta-analysis was published in 2010, ranging from one to six publications a year until 2017. In 2016 and 2017 systematic reviews and meta-analyses represented a third of all articles published on the topic (figure 5 and appendix 4).

**Figure 4:**
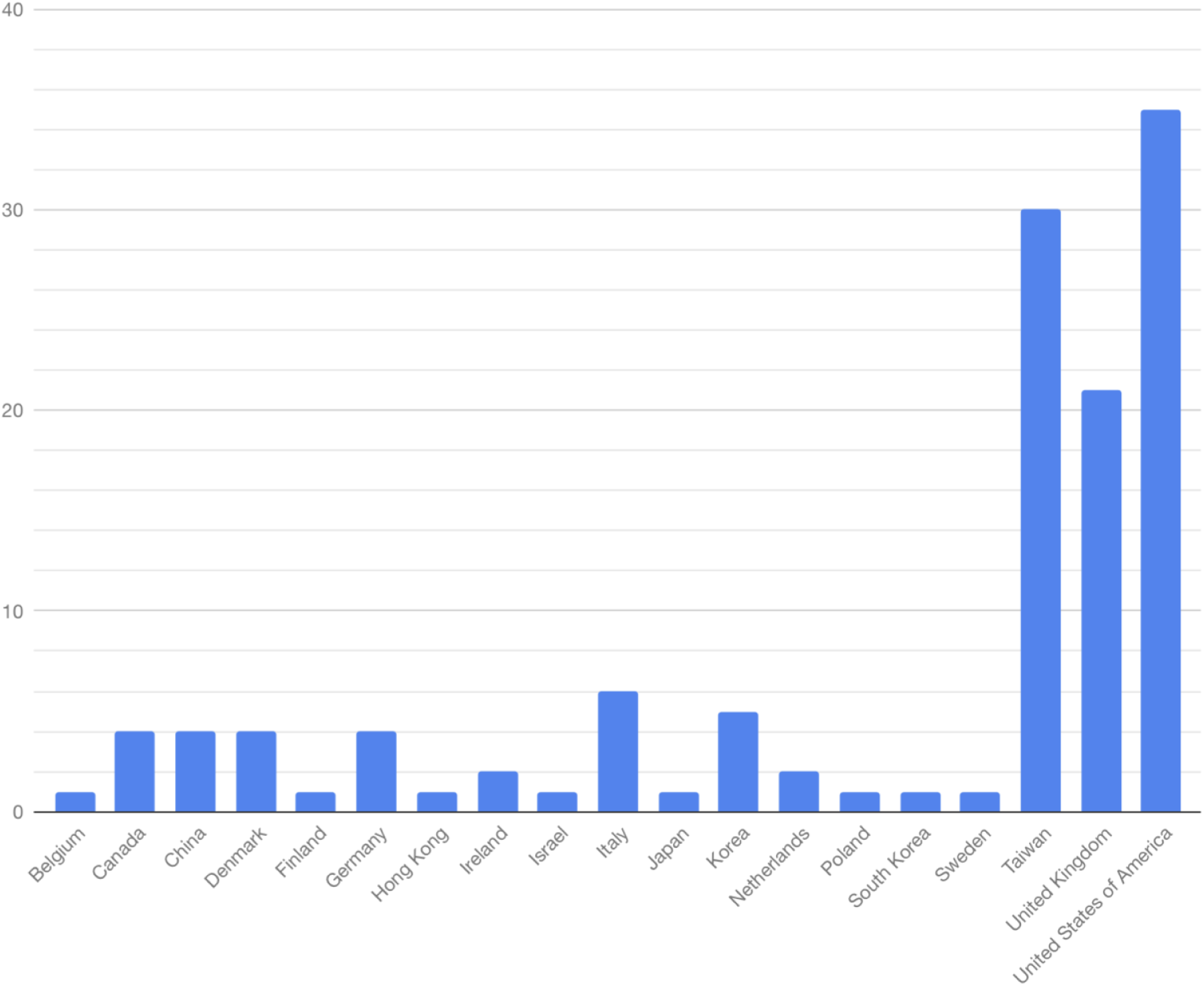
Included Studies by Country.

**Figure 5:**
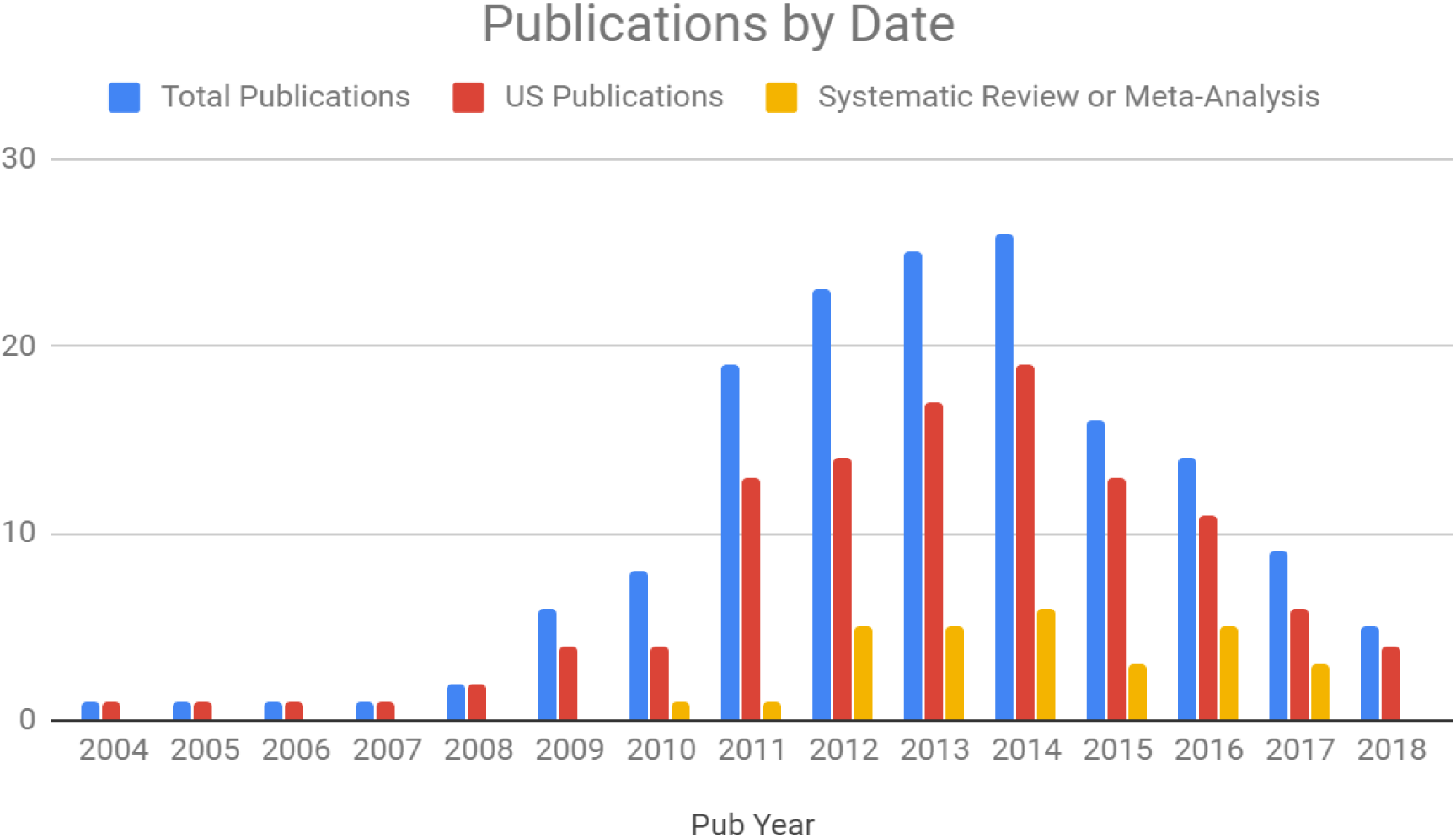
Total Publications, US Publication and Systematic Review or Meta-Analysis by date.

### Limitations

Due to the small number of authors available for this manuscript, elements of a rapid review were incorporated into the research methods^10^. Specifically, only subject headings were used in the PubMed search and not keywords. We believe this decision was justified as other searching techniques were used to offset this choice. The search used the subset cancer, which is a filter created by the National Library of Medicine and the National Cancer Institute, containing headings, keywords, and journal titles to facilitate searching for cancer. It contains over 1,000 terms in the search^174^. The subheading prevention and control was floated in the search, allowing it to attach itself to any term in the search, including the ones in the large cancer subset. Systematic Reviews and Meta-Analyses were included publications, so mining those articles for relevant references was also a strategy used to make a comprehensive and replicable search.

There were a couple of limitations in the analysis of the origins of the data. First, there were four articles that listed themselves as “international” studies but did not disclose what countries specifically. Those four articles were excluded from the country analysis ^38,61,108,168^. Also 20 studies listed themselves as taking place in the United Kingdom ^29–32,37,48,60,88,89,92–98,112,161,163,165,166^. The United Kingdom consists of four countries, England, Scotland, Wales and Northern Ireland. Since it could not be discerned which country within the United Kingdom it came from, the 20 studies were listed under the United Kingdom. Any studies that specified countries within the United Kingdom were also added to this group.Overall there was no data coming from African, Caribbean, Central and South American countries, which makes extrapolating results found in studies to those patient populations difficult.

There were limitations in the analysis of funding sources by cancer type. Many studies (40) listed the cancer type they were focusing on as “various” ^5,24,28–30,36,38,42,45,48–50,55–57,63,88–91,106,108,114,124,128,129,135,139,143,145,148,149,150,155,157,164,167,168,170,171^. It is complex to report the funding of these studies by cancer type because these were funded as multi-cancer studies. In addition, seven studies ^22,23,31,34,46,141,172^ did not specify what cancers they were focusing on which similarly made it difficult to analyze.

In the original protocol, there was the intent to identify the professions of the authors to map out which professions are actively participating in the research and publishing on this topic. However, most articles did not include that information and attempts to extract that data from websites produced inconsistent or non-existent data. During the mapping review process, information relating to metformin, such as dose and duration of use, were also considered. However, there were several variations of how these parameters were measured across the studies which made it difficult to analyze and report.

## Discussion

Metformin was first introduced into the USA in 1995. In 1998 the United Kingdom Prospective Diabetes Study (UKPDS) was published and it defined metformin’s place in therapy in the management of type 2 diabetes. Ten years later, a follow-up study was published and demonstrated continuous cardiovascular benefit of metformin when initiated early in the pharmacologic management of diabetes. In 2011, metformin was included in the essential medicines list published by the World Health Organization (WHO). This list is published every two years and presents minimum medication needs for a basic-health care system, and essential medicines for priority diseases^175^. This may explain the dramatic increase in the number of publications in 2012 and 2013.

The cancers represented in this review mirror those with the highest incidence in the United States. In 2019^176^ the cancers with the highest new cases were breast (#1 in female), prostate (#1 in male), lung (#2 in both genders) and colon cancer (#3 in both genders). This systematic mapping review shows that the most reviewed single cancers were colorectal (#1), prostate (#3), lung (#4) and breast (#5). The cancers that are in this review seem to mirror those most prevalent in the United States.

In 2010, the national cost of cancer care was highest for breast, colorectal, lymphoma, lung and prostate cancer respectively^177^. However, a similar study using data from 2007-2012, found that esophageal cancer was the second most expensive cancer at the initial phase and end of life care. Overall costs for cancer care annually were highest for esophageal at $25,508 for men and $23,007 for women. Additionally ovarian cancer had the highest cost for continuing care.^178^ Although the published literature of metformin and cancer has mirrored the incidence rates of cancer, perhaps more studies and funding are now needed to address the most costly cancers.

Metformin and its use to control or prevent cancer has the potential to be groundbreaking. Considering we can only extrapolate these results to populations that were represented in each of the studies some groups of people may be excluded from these findings. Those that would be included would be people from the represented countries stated in Figure 4, with cancers that were studied in abundance as shown in Figures 2 and 3. Results from this systematic mapping review may be less impactful in patients with rare cancers or from countries/regions that were not widely represented. This can be seen in the Caribbean as well as Central and South America, even though Spanish was included as a language in our search strategy.

The mean number of authors for the included systematic reviews and meta-analyses was one higher than the mean of five found by Borah in review of Prospero registries.^179^ Systematic reviews were also the lowest funded study types suggesting that this study design for metformin and cancer is more laborious than the global average, but not well supported financially, which might be a disincentive for authors to participate in this type of study.

Despite efforts to obtain large data sets for retrospective cohort studies in the United States, studies were limited by the segregated infrastructure of hospital systems and the difficulty of extracting and cleaning the meta-data from separate systems. Nation states that have a single payer healthcare system, like Taiwan, are able to analyze the entire population, not just segments, resulting in much more comprehensive data.

The review revealed no studies from the Caribbean, Central and South America despite Spanish being an inclusion criteria. Future research could build searches in the Spanish language and employ databases such as SciELO, Scientific Electronic Library Online, that caters to mostly Central and South American countries. Future reviews could also have no language limitations and contract out for translators to screen and extract data.

## Conclusion

Type 2 diabetes is a chronic disease state that has been known to increase the risk of contracting various cancers. Metformin as the first line treatment has made its mark on the diabetes community. Not only is it preferred due to effectiveness, cost, and side effect profile it has also been extensively researched in its impact in the prevention and control of cancer. This systematic mapping review categorizes this literature. A total of 156 articles were included in the analysis.

Colorectal, liver, prostate, lung, and breast cancer were the most researched and most funded cancers. A majority of studies included received at least one source of funding. The country with the most literature published was the United States. Only 68% of all studies were available open access. Especially since the number of publications has decreased annually since 2014, future research efforts and funding need to be judicious and address not only mortality and prevalence, but total cancer cost as well as underrepresentation when choosing a cancer to pursue. Additionally, those practitioners working with underrepresented cancers as identified in this systematic mapping review, have to rely on scarce data or indirect evidence from other studies, negating their ability to use evidenced-based care regarding metformin and cancer to their patients.

## Supporting information

PRISMA 2020 Checklist

Cipriano COI

Mitchell COI

Price COI

## Data Availability

The data is available in the tables or appendices.

## Author Contributions

- Designed the Review/Conceived the Review-MP
- Built Search Strategy - MP
- Literature Extraction - MP
- Screened Titles - GC, AM
- Extracted Data - GC, AM, MP
- Wrote First Draft/Drafted the Manuscript - GC, AM, MP
- Researched & Interpreted Data - GC, AM, MP
- Reviewed Draft/Manuscript Revision- GC,AM, MP
- Reviewed Final manuscript for Intellectual Content-GC, AM, MP
- Approved Final Draft -GC,AM,MP

## Appendix 1

Search Strategy for PubMed

1. Metformin[MeSH]
2. “Diabetes Mellitus, Type 2”[MeSH]
3. “prevention and control”[sh]
4. cancer[sb]
5. #1 AND #2 AND #3 AND #4

Search Strategy for Cochrane Library

1. MeSH Descriptor: [Metformin] explode all trees
2. Metformin
3. #1 OR #2
4. MeSH Descriptor: [Diabetes Mellitus, Type 2] explode all trees
5. MeSH Descriptor: [Neoplasms] explode all trees
6. Cancer
7. #5 OR #6
8. #3 AND #4 AND #7

Search Strategy for ClinicalTrials.gov

Condition or Disease: Diabetes Mellitus, Type 2

Other Terms: metformin and cancer

## Appendix 2 Funders

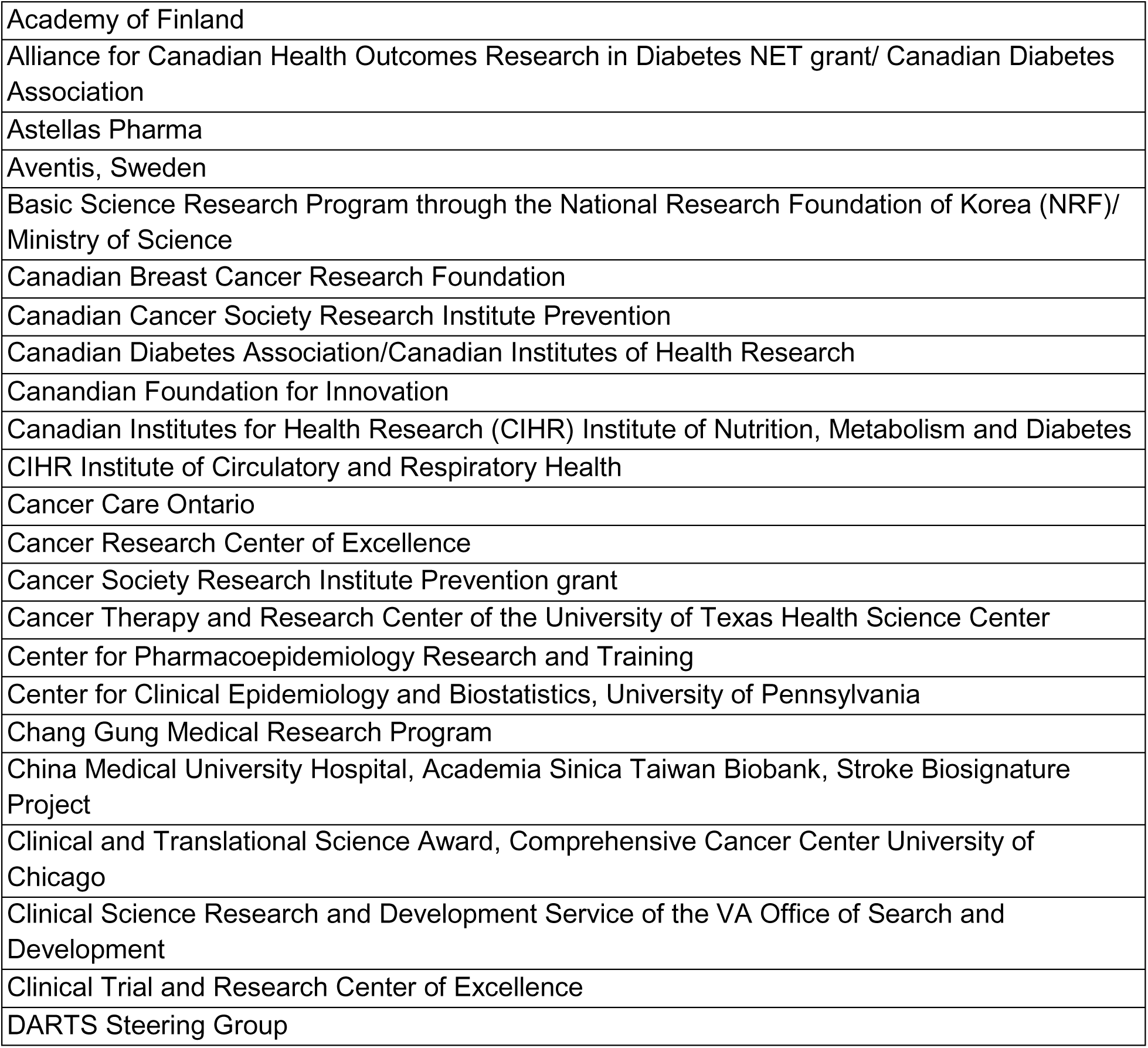
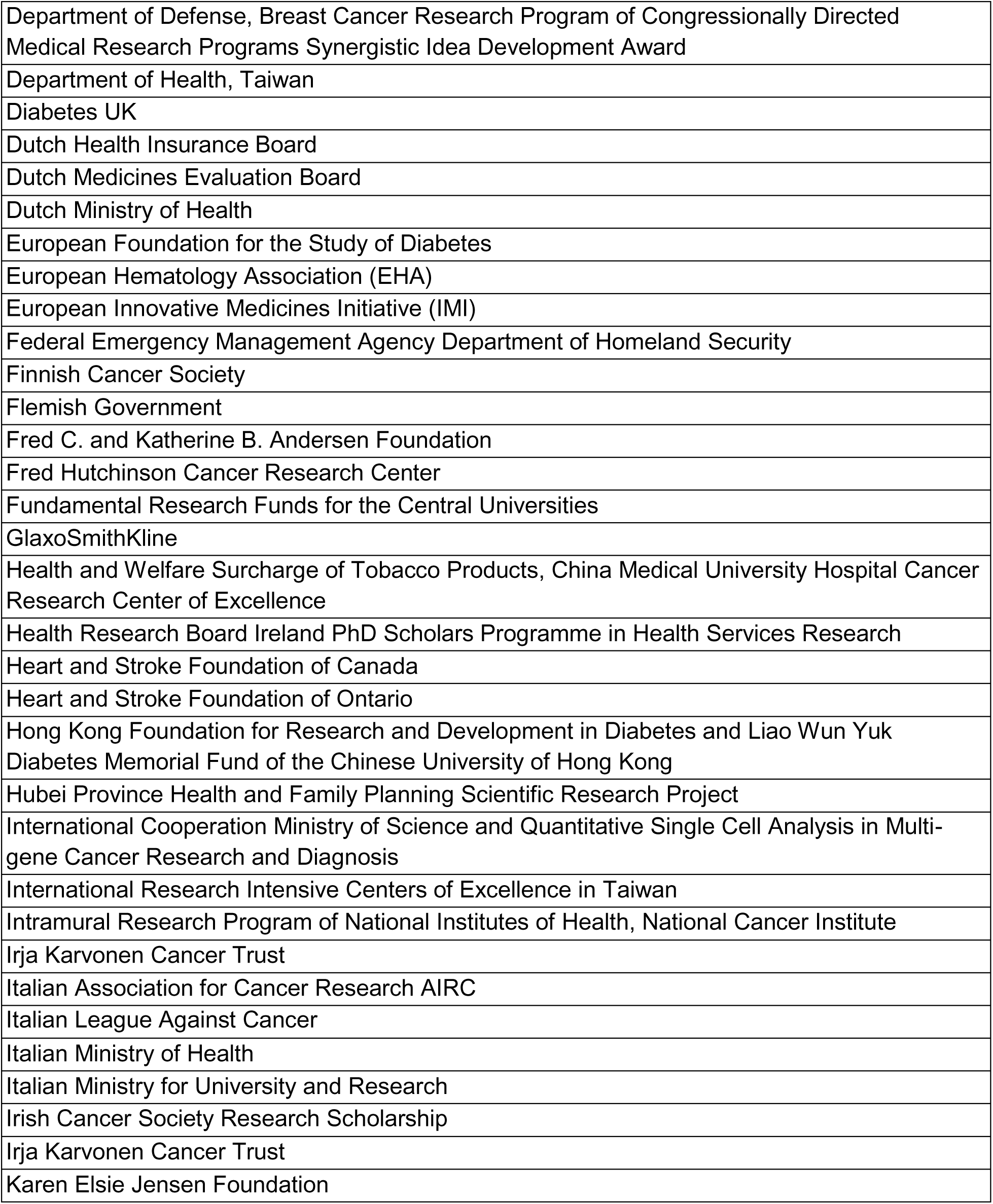
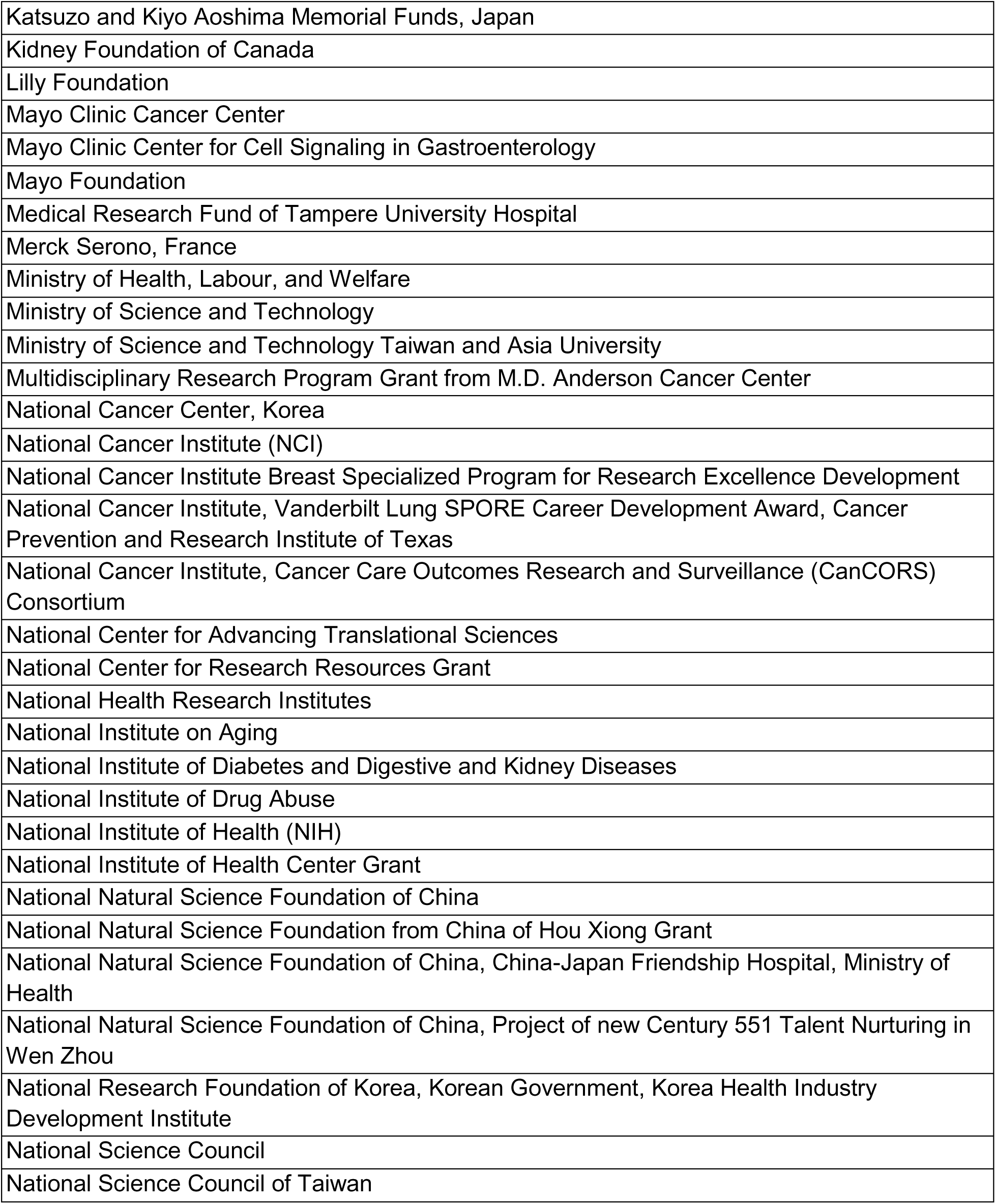
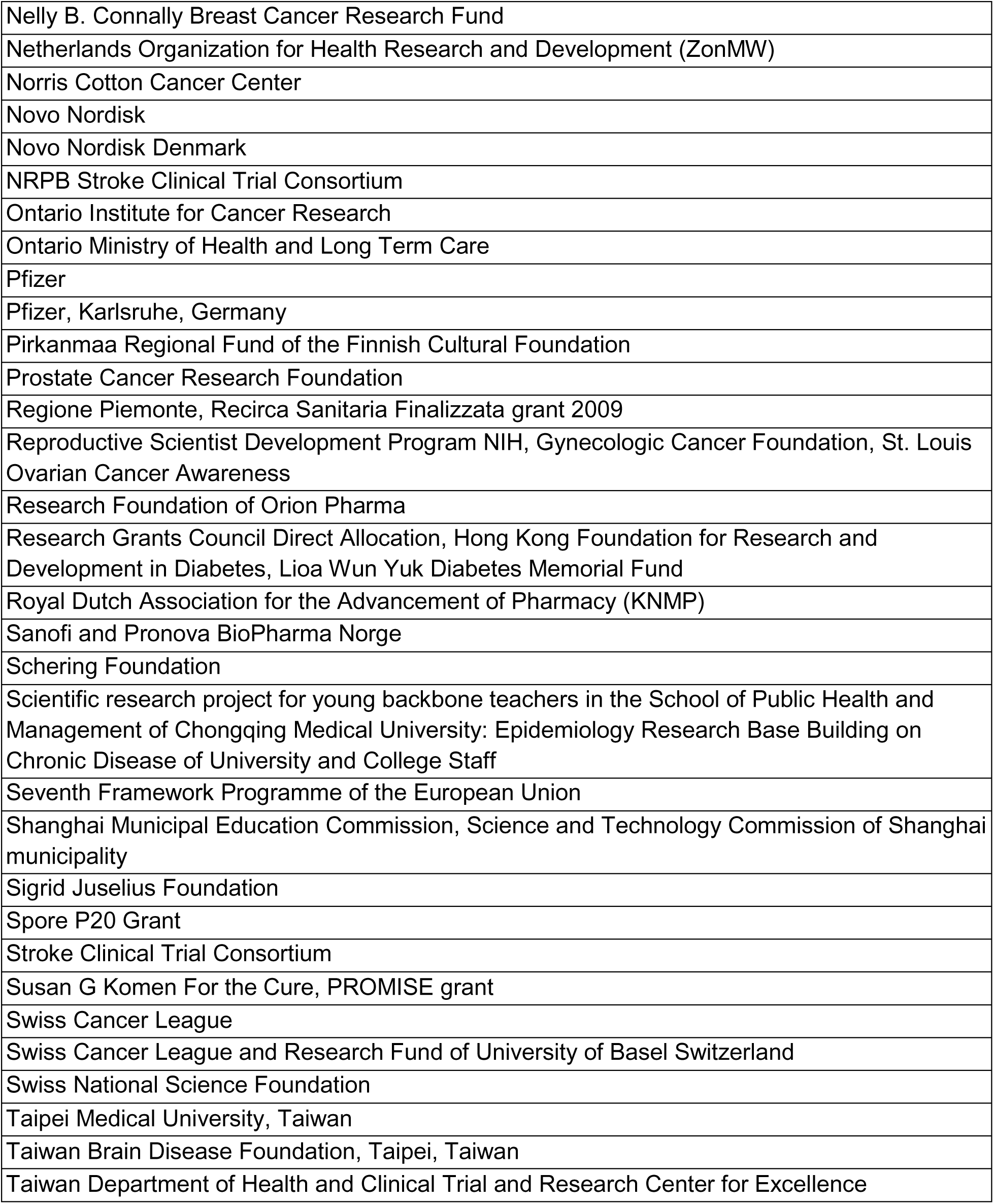
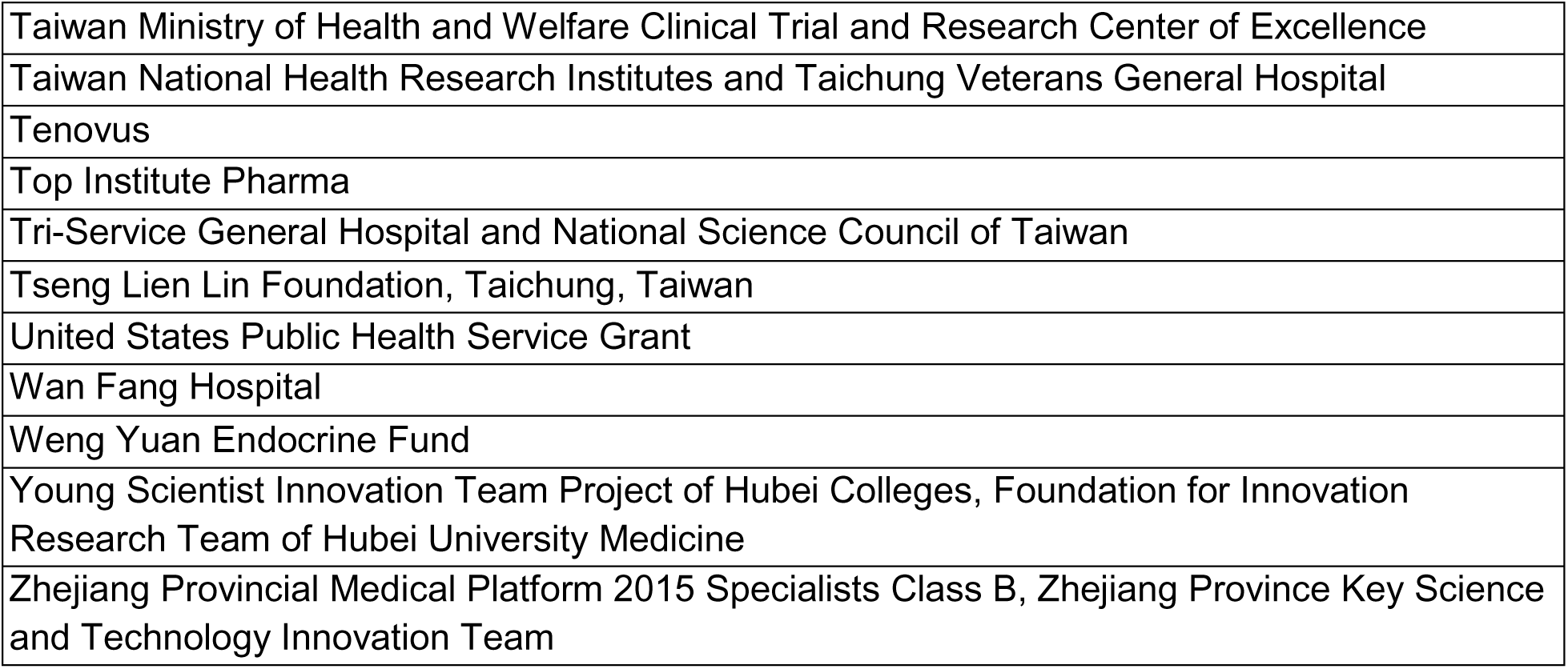

## Appendix 3 Articles by Journal Title

| Number of articles | Journal |
| --- | --- |
| 16 | Diabetes Care |
| 8 | Cancer Epidemiology, Biomarkers and Prevention |
| 7 | Cancer |
| 7 | Oncotarget |
| 5 | Diabetologia |
| 4 | Gynecologic Oncology; PlosOne; Metabolism: Clinical and Experimental; Medicine |
| 3 | American Journal of Gastroenterology; Breast Cancer Research and Treatment; Cancer Prevention Research; Livert International |
| 2 | Acta Diabetologica; American Journal of Epidemiology; Annals of Oncology; Asian Pacific Journal of Cancer Prevention; BMC Cancer; British Journal of Cancer; Cancer Causes and Control; Cancer Epidemiology; Current Drug Safety; Diabetes/Metabolism Research and Reviews; Diabetes Research and Clinical Practice; European Journal of Endocrinology; Gastroenterology; Gut; Journal of Clinical Endocrinology and Metabolism; Journal of Clinical Oncology; Lung Cancer; Oncologist; World Journal of Gastroenterology |
| 1 | Aging; Alimentary Pharmacology & Therapeutics; The American Journal of the Medical Sciences; Anti-Cancer Research; Biomedical Reports; BMC Medicine; BMJ; Cancer Medicine; Clinical Cancer Research; Clinical Lung Cancer; Diabetologia and Metabolic Syndrome; Diabetes Complications; Diabetes & Metabolism Journal; Diabetes Obesity and Metabolism; Digestive and Liver Disease; Endocrinologia Polyska; Epidemiology/Health Services Research; European Journal of Internal Medicine; European Urology; Experimental Diabetes Research; Experiment and Clinical Endocrinology and Diabetes; Head and Neck; Hepatology; International Journal of Cancer; Intestinal Research; Journal of the American Academy of Dermatology; Journal of the American Medical Informatics; Journal of Diabetes and its |
|  | Complication; Journal of Gastroenterology and Hepatology; Journal of Gastrointestinal Cancer; The Journal of Thoracic and Cardiovascular Surgery; Journal of the National Cancer Institute; Lancet Oncology; Medical Care; Obstetrics and Gynecology; Pharmaceutical Biology; Pharmacoepidemiology and Drug Safety; Pharmacotherapy Publications; Prostate Cancer and Prostatic Disease; Scandinavian Journal of Gastroenterology; Scientific Reports; Southern Medical Association Journal; Therapeutics and Clinical Risk Management; Urologic Oncology; Urology; World Journal of Urology |

## Appendix 4 Studies Organized by Publication Date

| Publication Year | 2004 | 2005 | 2006 | 2007 | 2008 | 2009 | 2010 | 2011 | 2012 | 2013 | 2014 | 2015 | 2016 | 2017 | 2018 |
| --- | --- | --- | --- | --- | --- | --- | --- | --- | --- | --- | --- | --- | --- | --- | --- |
| Total Publications | 1 | 1 | 1 | 1 | 2 | 6 | 8 | 19 | 23 | 25 | 26 | 16 | 14 | 9 | 5 |
| US Publications | 1 | 1 | 1 | 1 | 2 | 4 | 4 | 13 | 14 | 17 | 19 | 13 | 11 | 6 | 4 |
| Systematic Review or Meta-Analysis | 0 | 0 | 0 | 0 | 0 | 0 | 1 | 1 | 5 | 5 | 6 | 3 | 5 | 3 | 0 |
| US Publications Percent of Total | 100% | 100% | 100% | 100% | 100% | 67% | 50% | 68% | 61% | 68% | 73% | 81% | 79% | 67% | 80% |
| Systematic Review or Meta-Analysis Percent of Total | 0% | 0% | 0% | 0% | 0% | 0% | 13% | 5% | 22% | 20% | 23% | 19% | 36% | 33% | 0% |

